# Fluvoxamine for Outpatient Treatment of COVID-19: A Decentralized, Placebo-controlled, Randomized, Platform Clinical Trial

**DOI:** 10.1101/2022.10.17.22281178

**Authors:** Matthew W. McCarthy, Susanna Naggie, David R. Boulware, Christopher J. Lindsell, Thomas G. Stewart, G. Michael Felker, Dushyantha Jayaweera, Mark Sulkowski, Nina Gentile, Carolyn Bramante, Upinder Singh, Rowena J. Dolor, Juan Ruiz-Unger, Sybil Wilson, Allison DeLong, April Remaly, Rhonda Wilder, Sean Collins, Sarah E. Dunsmore, Stacey J. Adam, Florence Thicklin, George Hanna, Adit A. Ginde, Mario Castro, Kathleen McTigue, Elizabeth Shenkman, Adrian F. Hernandez, the Accelerating COVID-19 Therapeutic Interventions and Vaccines (ACTIV)-6 Study Group

**Author notes:** **Trial Registration:** ClinicalTrials.gov (NCT04885530).

## Abstract

**Background:** The effectiveness of fluvoxamine to shorten symptom duration or prevent hospitalization among outpatients in the US with mild to moderate symptomatic coronavirus disease 2019 (COVID-19) is unclear.

**Design:** ACTIV-6 is an ongoing, decentralized, double-blind, randomized, placebo-controlled platform trial testing repurposed medications in outpatients with mild to moderate COVID-19. A total of 1288 non-hospitalized adults aged ≥30 years with confirmed COVID-19 experiencing ≥2 symptoms of acute infection for ≤7 days prior to randomization were randomized to receive fluvoxamine 50 mg or placebo twice daily for 10 days. The primary outcome was time to sustained recovery, defined as the third of 3 consecutive days without symptoms. Secondary outcomes included composites of hospitalization or death with or without urgent or emergency care visit by day 28.

**Results:** Of 1331 participants randomized (mean [SD] age, 48.5 [12.8] years; 57% women; 67% reported receiving at least 2 doses of a SARS-CoV-2 vaccine), 1288 completed the trial (n=614 placebo, n=674 fluvoxamine). Median time to recovery was 13 days (IQR 12–13) in the placebo group and 12 days (IQR 11–14) in the fluvoxamine group (hazard ratio [HR] 0.96, 95% credible interval [CrI] 0.86–1.07; posterior probability for benefit [HR>1]=0.22). Twenty-six participants (3.9%) in the fluvoxamine group were hospitalized or had urgent or emergency care visits compared with 23 (3.8%) in the placebo group (HR 1.1, 95% CrI 0.6–1.8; posterior probability for benefit [HR<1]=0.340). One participant in the fluvoxamine group and 2 in the placebo group were hospitalized; no deaths occurred. Adverse events were uncommon in both groups.

**Conclusions:** Treatment with fluvoxamine 50 mg twice daily for 10 days did not improve time to recovery, compared with placebo, among outpatients with mild to moderate COVID-19. These findings do not support the use of fluvoxamine at this dose and duration in patients with mild to moderate COVID-19.

## INTRODUCTION

There remains a need for oral therapies to prevent progression to severe coronavirus disease 2019 (COVID-19).^1^ Novel oral antivirals have demonstrated a clinical benefit in unvaccinated persons; however, the efficacy of current recommended therapies for mild to moderate COVID-19 for vaccinated patients is unclear and may be lower than what was reported in unvaccinated populations.^2,3^ Repurposing approved drugs developed for other conditions presents an attractive strategy for the identification of new treatment options and expanding access to potentially life-saving care.^4,5^

Fluvoxamine is a selective serotonin reuptake inhibitor approved by the U.S. Food and Drug Administration (FDA) in 1994 for the treatment of obsessive-compulsive disorder and is now used to treat a variety of psychiatric conditions, including social anxiety disorder and depression.^6^ Fluvoxamine has also been noted to activate the sigma-1 receptor, which may decrease inflammation by reducing endoplasmic reticulum stress and down-regulating the expression of inflammatory genes. Early studies in patients with COVID-19 reported improved clinical outcomes in participants receiving fluvoxamine.^7-9^

A placebo-controlled, randomized, adaptive platform (TOGETHER) trial of 1497 symptomatic Brazilian adults with confirmed severe acute respiratory syndrome coronavirus 2 (SARS-CoV-2) and a known risk factor for progression to severe COVID-19 found treatment with fluvoxamine 100 mg twice daily for 10 days reduced the need for hospitalization, defined as retention in an emergency setting or transfer to a tertiary hospital.^10^ However, tolerability has been identified as a potential limiting factor for this dose of fluvoxamine, with 74% of participants in the fluvoxamine group reporting completion of more than 80% of possible doses. The STOP COVID 2 trial, in which participants were randomized to receive 100 mg of fluvoxamine twice daily or placebo for 15 days, was stopped for futility in May 2021 after an interim analysis found that the low event rate seen in the trial with the original sample size was associated with a less than 10% conditional probability of demonstrating efficacy of fluvoxamine.^11^ A subsequent meta-analysis of 3 clinical trials found a high probability (94.1–98.6%) that fluvoxamine was associated with a reduced risk for hospitalization from COVID-19, with a risk ratio of 0.75 (95% confidence interval [CI] 0.58–0.97).^7^ Given these conflicting results, regulatory authorities and guideline committees have not recommended fluvoxamine as early treatment for COVID-19. Uncontrolled observational data suggested a lower 50 mg dose would be more tolerable.^12^

We sought to investigate a 50 mg twice daily dose of fluvoxamine for 10 days in a double-blind, randomized, placebo-controlled, platform trial investigating repurposed drugs for non-hospitalized persons with mild to moderate COVID-19.

## METHODS

### Trial Design and Oversight

Accelerating COVID-19 Therapeutic Interventions and Vaccines (ACTIV)-6 (NCT04885530) is a double-blind, randomized, placebo-controlled platform protocol conducted using a decentralized approach. ACTIV-6 enrolls outpatients with mild to moderate COVID-19 with a confirmed positive polymerase chain reaction or antigen test for SARS-CoV-2 infection, including home-based testing.

The protocol was approved by each site’s institutional review board. Informed consent was obtained from each participant either via written or electronic consent. An independent data monitoring committee oversaw participant safety and trial performance.

### Participants

Recruitment into the platform trial opened on June 11, 2021 and is ongoing. Participants were enrolled into the fluvoxamine group or matched or contributing placebo from August 6, 2021 through May 27, 2022 at 91 sites in the United States. Participants were identified by trial sites or they self-identified either by registering online or by contacting a central study telephone hotline. Participants without a local site were managed by the coordinating center call center.

Sites verified eligibility criteria including age ≥30 years, confirmed SARS-CoV-2 infection ≤10 days, and experiencing ≥2 COVID-19 symptoms for ≤7 days from time of consent. Symptoms included the following: fatigue, dyspnea, fever, cough, nausea, vomiting, diarrhea, body aches, chills, headache, sore throat, nasal symptoms, and new loss of sense of taste or smell. Exclusion criteria included hospitalization, known allergy or contraindication including prohibited concomitant medications, or study drug use within 14 days of enrollment (see **Online Supplement** for full eligibility criteria). Vaccination was not an exclusion. Standard COVID-19 therapies available under FDA approval or emergency use authorization were allowable.

### Randomization and Interventions

Within the platform trial, study drugs could be added or removed according to adaptive design and/or emerging evidence. Participants were randomized to one of the study drugs actively enrolling at the time of randomization, which included oral ivermectin 400 μg/kg/day for 3 days or inhaled fluticasone furoate 200 μg once daily for 14 days or placebo during the period that the fluvoxamine group was open. As multiple study drugs were available, randomization occurred based on appropriateness of each drug for the participant as determined by the eligibility criteria. Participants could choose to opt out of specific groups during the consent process.

ACTIV-6 was designed to share information from participants randomized to receive placebo. At the first step of randomization, participants were assigned to receive either placebo or active drug in the ratio 1:*m*, where *m* is the number of study drug groups for which participants were eligible. Subsequently, participants are randomized among the *m* study groups with equal probability. Participants randomized to receive placebo contribute to all of the study groups for which eligibility was met.

Participants received study drug via direct home delivery from a central pharmacy with a 10-day supply of either fluvoxamine in a foil blister strip or identical matched placebo foil blister strip, provided by the manufacturer (Abbott Laboratories, Abbott Park, IL). Participants self-administered 50 mg (1 blister) of fluvoxamine or matching placebo twice daily for 10 days.

### Outcome Measures

The primary measure of effectiveness was time to recovery, defined as the third of 3 consecutive days without symptoms. This was selected *a priori* from among the 2 co-primary outcomes that remain available to other platform study drugs. The key secondary outcome was hospitalization or death by day 28. Other secondary outcomes included time unwell; COVID-19 Clinical Progression Scale on days 7, 14, and 28; mortality through day 28; and urgent or emergency care visit or hospitalization through day 28.

### Trial Procedures

ACTIV-6 is a decentralized trial; thus, all study visits are planned as remote. Screening and eligibility confirmation are participant-reported and confirmed by trial sites. A positive SARS-CoV-2 test result was verified by sites prior to randomization. At the screening visit, participants reported demographic information, eligibility criteria, medical history, concomitant medications, symptoms, and completed quality of life questionnaires.

A central investigational pharmacy distributed study drug to residential addresses provided by participants, and shipping and delivery were tracked. Participants must have received study drug to be included in the analysis. Receipt of study drug was defined as day 1.

Participants were asked to complete assessments and report safety events daily through the first 14 days of study. From days 15–28, participants continued to report if they had symptoms until they had experienced 3 consecutive days without symptoms. Follow-up visits occurred at day 28 and day 90. At each study assessment, participants self-reported symptoms and severity, health care visits, and any new medications.

The daily and follow-up assessments were monitored, and sites were actively notified of events requiring review, including serious adverse events (SAEs). In addition, participants were invited during assessments to request contact from the study team or to report any unusual circumstances. Failure to complete daily assessments also triggered a review for any possible SAEs. A missed assessment on the day after receiving the first dose of study medication (day 2) or any day of missed assessments up to day 14 prompted a notification to the site to contact the participant. All participants were instructed to self-report concerns either via an online event reporting system, by calling the site, or by calling a 24-hour hotline.

### Statistical Analysis Plan

The ACTIV-6 trial is designed to be analyzed using a Bayesian approach. Decision thresholds were set to balance overall power with control of the Type I error rate in the context of a study drug-specific goal. An estimated sample size of approximately 1200 participants was expected to be sufficient to conclude whether there is meaningful evidence of clinical benefit on the primary outcome.

As a platform trial, the primary analysis is implemented separately for each study drug, where the placebo group consists of concurrently randomized participants who met enrollment criteria for that study drug, including consent to be randomized to fluvoxamine. A modified intention-to-treat (mITT) approach was specified for primary analyses, including all participants who received study drug. From other remote trials,^13,14^ it was recognized that medication delivery (placebo or study drug) may not always occur (e.g., failure of delivery, participant withdrawal, or interval hospitalization). This resulted in exclusion of the participant for the mITT analysis. All available data were utilized to compare fluvoxamine versus concurrent placebo control, regardless of post-randomization adherence. The safety population included those participants in the mITT population who reported taking at least 1 dose of study drug or matching placebo.

Heterogeneity of treatment effect was assessed for preselected subgroups and for the following covariates: age, sex, duration of symptoms, body mass index (BMI), symptom severity, calendar time (corresponding to predominant SARS-CoV-2 variant), and vaccination status.

## RESULTS

Of the 3837 participants who consented to be evaluated for inclusion in the fluvoxamine group, 1331 were randomized to fluvoxamine 50 mg twice daily (n=686) or placebo (n=645), and there were n=674 participants receiving study drug in the fluvoxamine group, and n=614 receiving study drug in the placebo group (**Figure 1**). Of participants receiving placebo, 326 (53%) received matching placebo and 288 (47%) contributed from the pooled placebo group.

**Figure 1.**
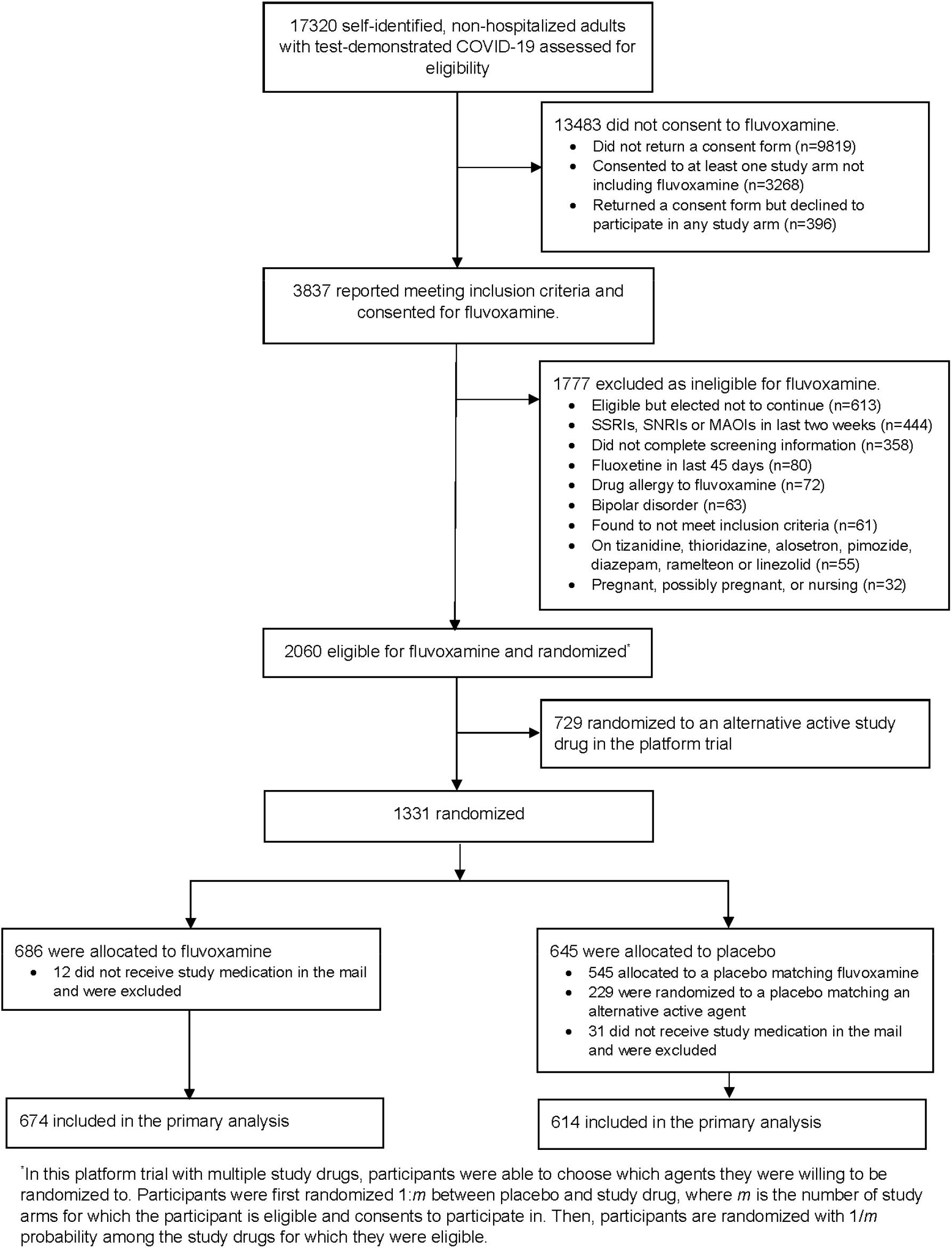
CONSORT diagram.

The mean (SD) age of the participants was 48.5 (12.8) years, and 42% were ≥50 years of age (**Table 1**). The population was 57% female, 81% identified as White, 7.5% Black/African American, 6.4% Asian, and 17% Latino/Hispanic ethnicity. Although not required for enrollment, high risk co-morbidities were prevalent, including BMI >30 kg/m^2^ (36.4%), diabetes (9.2%), hypertension (24.4%), and asthma (13.2%). Vaccination was common, with 67% of participants reporting at least 2 doses of a SARS-CoV-2 vaccine. The median time from symptom onset to receipt of study drug was 5 days (interquartile range [IQR] 4–7) with 77% and 80% receiving placebo and study drug, respectively, within 7 days of symptom onset (**eFigure 1**). Baseline symptom prevalence and severity are described in **eTable 1**. Although allowable per protocol, therapeutics available under FDA approval or authorization were uncommonly used (remdesivir 0.08%, monoclonal antibody 1.6%, ritonavir-boosted nirmatrelvir 1%) (**Table 1**).

**Table 1.**
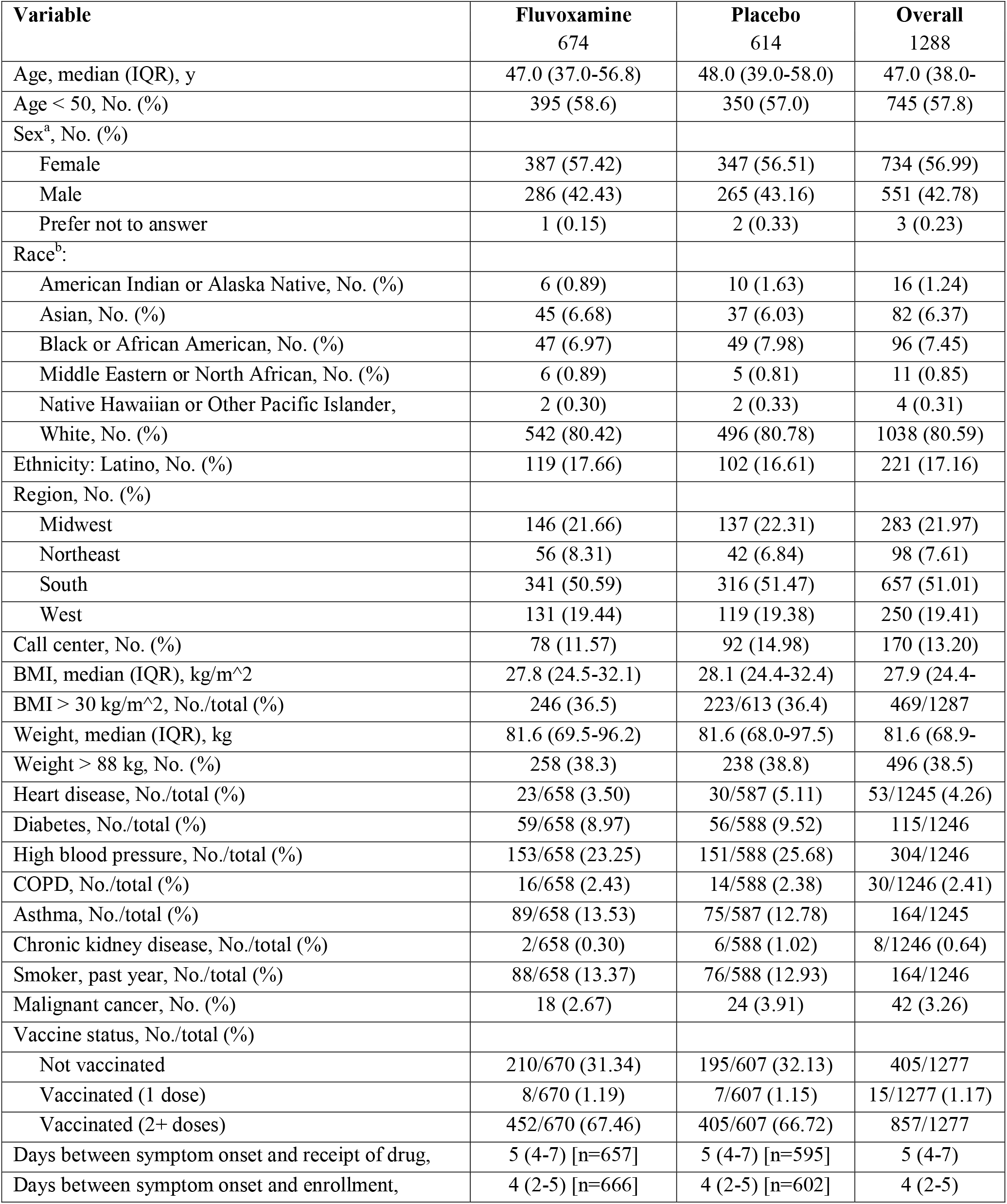
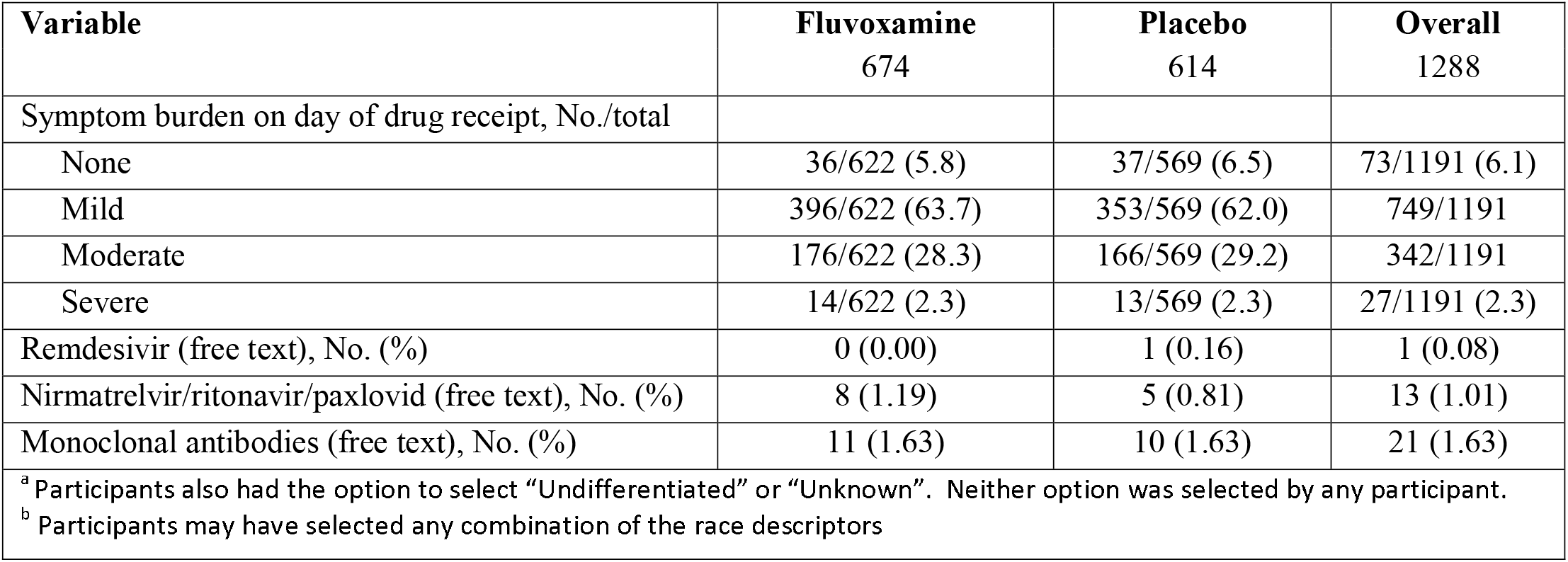
Baseline characteristics

### Primary Outcome

In the mITT population, the posterior probability for benefit on the primary outcome of time to recovery between the fluvoxamine and placebo groups was 0.22 (hazard ratio [HR] 0.96, 95% credible interval [CrI] 0.87–1.07) where a HR>1 indicates faster symptom resolution in the active drug group (**Table 2, Figure 2A**). The median time to recovery was 12 days (IQR11–14) in the fluvoxamine group and 13 days (IQR 12–13) in the placebo group. There remained no statistical evidence of a treatment benefit when using a non-informative prior, no prior, and with various approaches to imputing missing symptom data (**Figure 3**).

**Table 2.** Primary and secondary outcomes

| Endpoint | Fluvoxamine | Placebo | Estimate<br>(95% Interval) | Posterior<br>P(efficacy) |
| --- | --- | --- | --- | --- |
| Time to recovery |  |  |  |  |
| Skeptical prior (primary) |  |  | HR: 0.96 (0.87, 1.07) | 0.22 |
| Non-informative prior |  |  | HR: 0.94 (0.82, 1.06) | 0.17 |
| No prior |  |  | HR: 0.94 (0.83, 1.07) | • |
| Mean time unwell, days (95% CrI) | 11.22 (11.00, 11.41) | 11.27 (11.08, 11.47) | $\Delta$: -0.06 (-0.43, 0.33) | 0.613 |
| Mortality (day 28), No./total (%) | 0/662 (0.00) | 0/599 (0.00) | • | • |
| Hospitalization or death through day 28, No./total (%) | 1/670 (0.15) | 2/607 (0.33) | HR: 0.45 (0.04, 4.99) | • |
| Hospitalization, urgent care, emergency room visit, or death through day 28, No./total (%) | 26/670 (3.88) | 23/607 (3.79) | HR: 1.1 (0.6, 1.8) | 0.340 |
| Clinical progression ordinal outcome scale |  |  |  |  |
| Day 7 |  |  | OR: 1.28 (0.69, 1.88) | 0.173 |
| Day 14 |  |  | OR: 1.17 (0.49, 2.01) | 0.386 |
| Day 28 |  |  | OR: 1.46 (0.79, 2.28) | 0.103 |

Dots indicate not evaluable.
OR>1 favors placebo.

**Figure 2.**
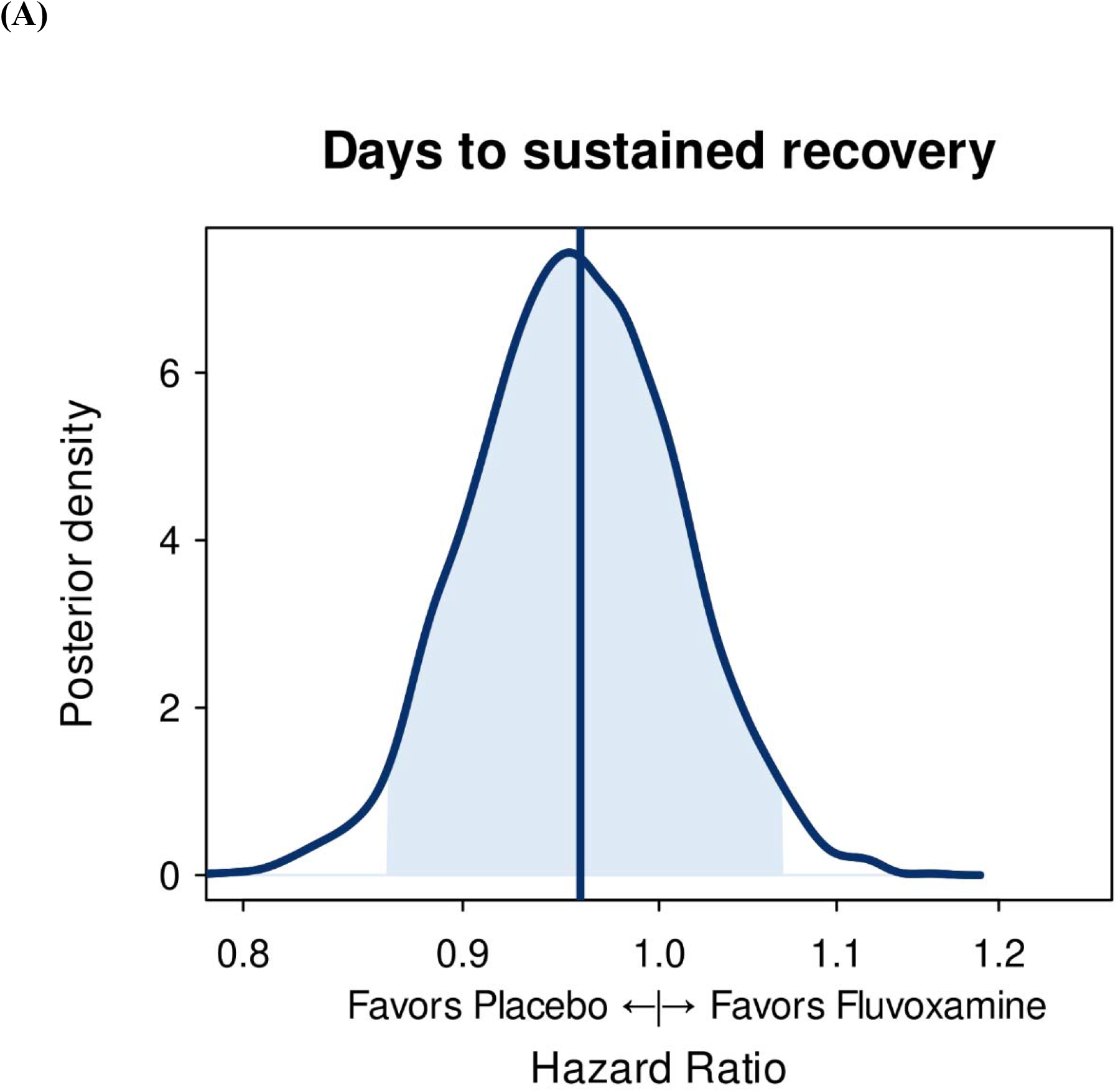
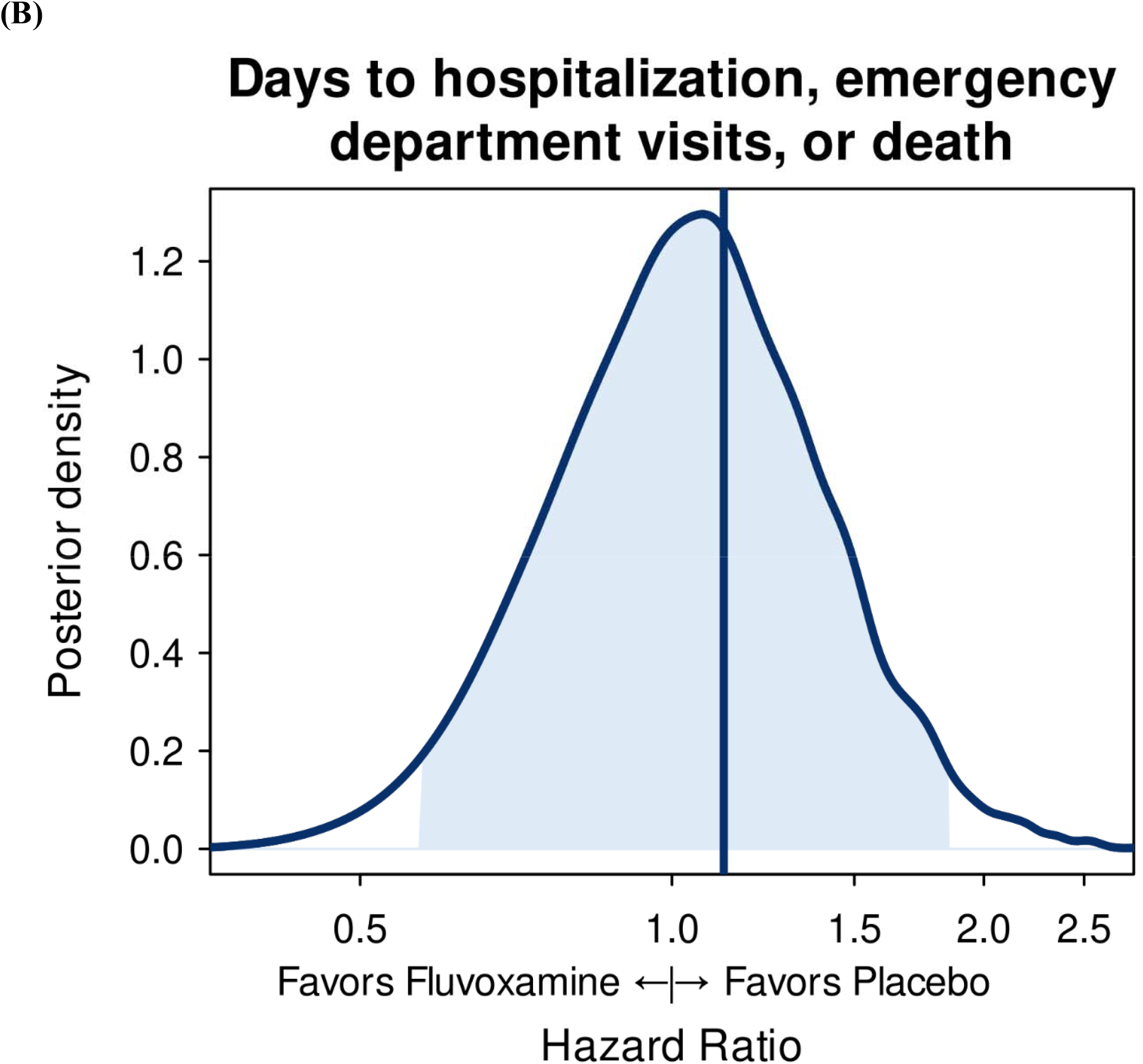
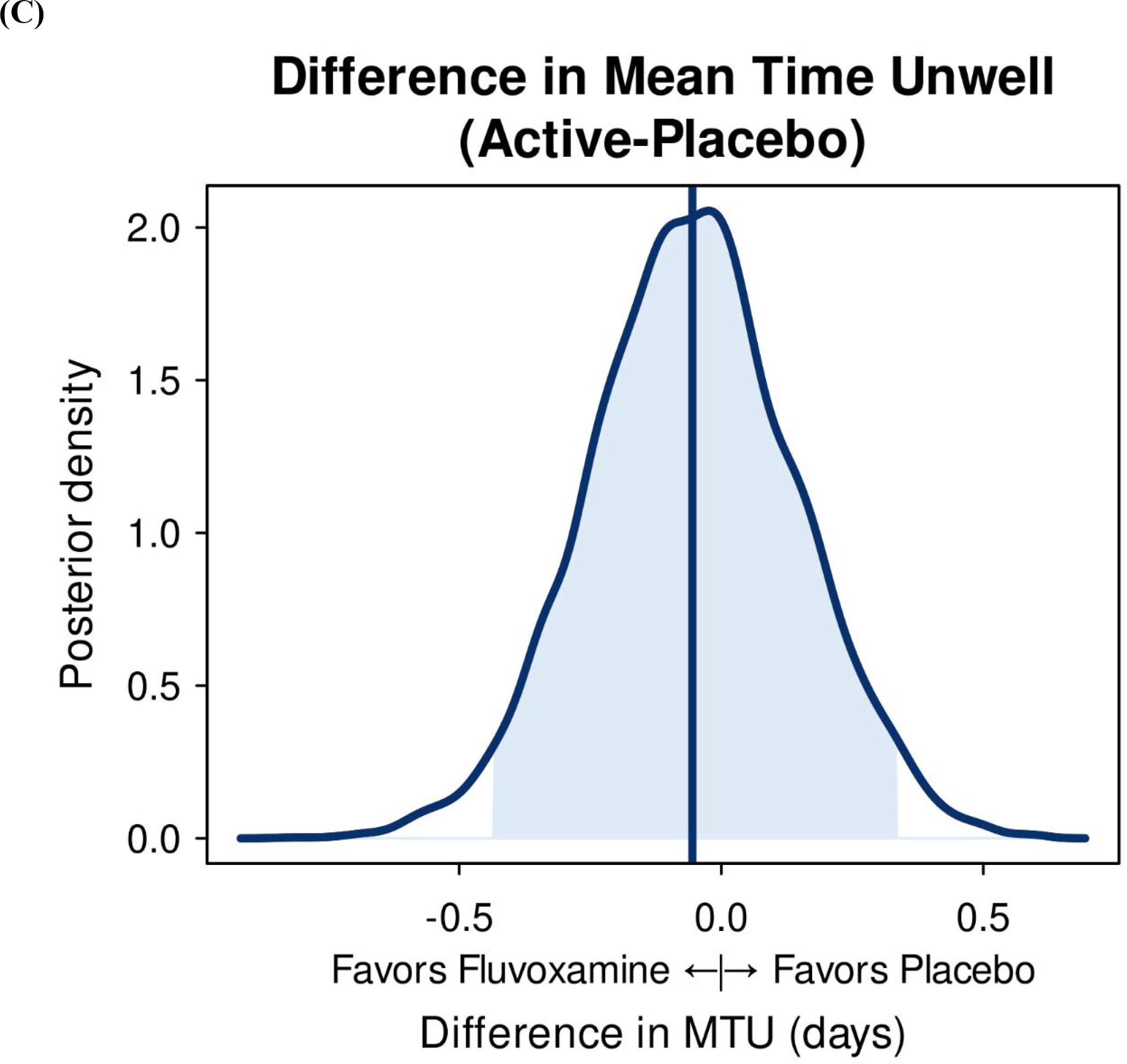
Posterior distributions of effects of (A) time to sustained recovery; (B) Hospitalization, urgent or emergency care visits, or death; (C) mean time unwell.

**Figure 3.**
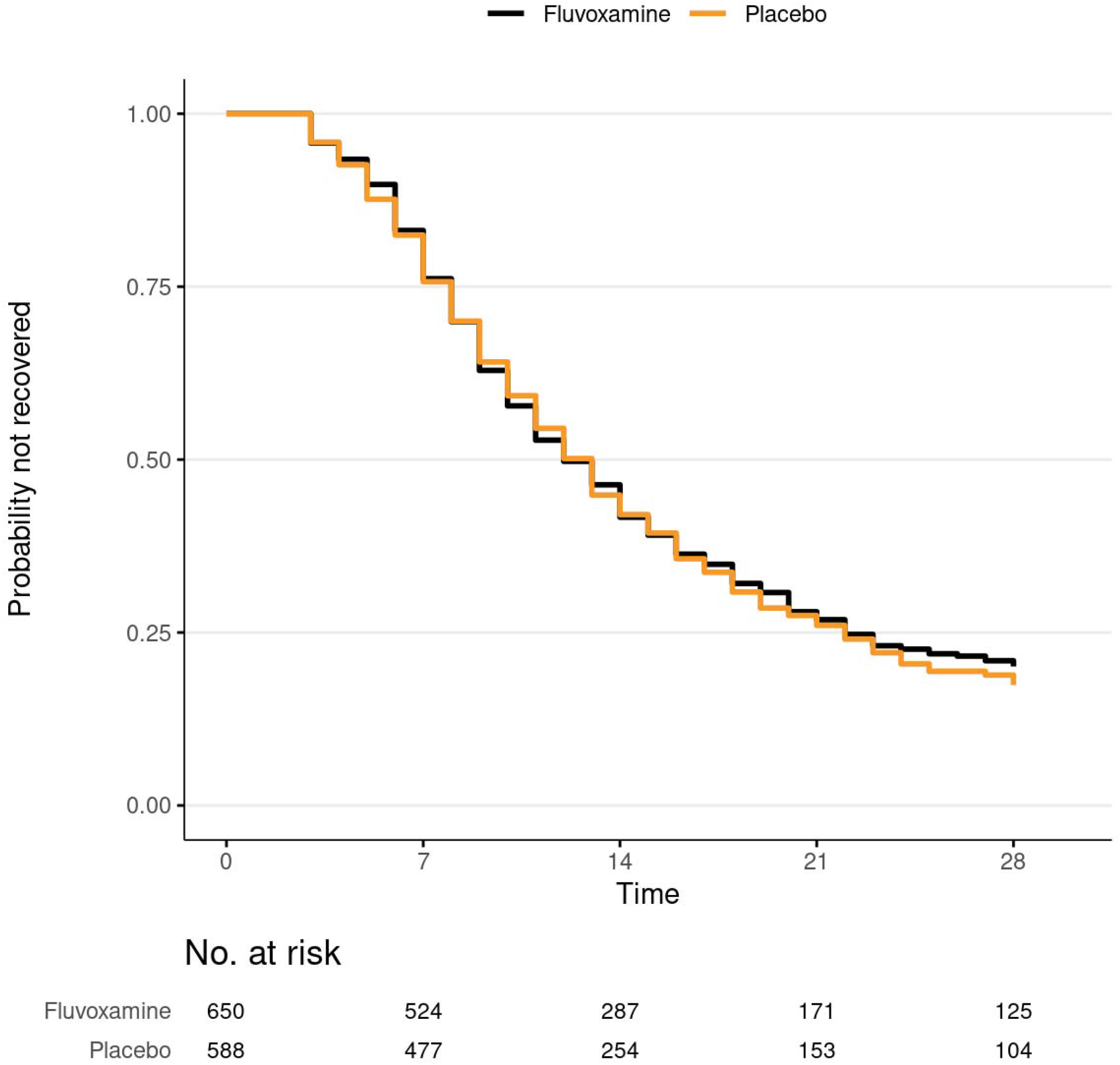
Kaplan-Meier for primary outcome of time to sustained recovery.

### Secondary Outcomes

Hospitalizations were uncommon, with 1 in the fluvoxamine group and 2 in the placebo group, and there were no deaths in the study (**eFigure 2A**). Thus, the HR on this composite outcome was uninformative. The composite secondary outcome of urgent or emergency care visits, hospitalizations, or death was similar with fluvoxamine (3.9% [26/670]) compared with placebo (3.8% [23/607]) (HR, 1.1 [95% CrI 0.6–1.8] where an HR>1favors placebo) (**Table 2, Figure 2B, eFigure 2B**). The difference in the amount of time spent feeling unwell with COVID-19 was 0.06 days (95% CrI -0.33 to 0.43 days) in favor of placebo (**Figure 2C**). The posterior probability of any benefit observed with the COVID Clinical Progression Scale (**Online Supplement**) at days 7, 14, and 28 also did not meet prespecified thresholds for beneficial treatment effect. For example, by day 7, 93.1% (604/649) of the fluvoxamine group and 93.9% (559/595) of the placebo group were not hospitalized and did not report limitation of activities (**eFigure 3**).

### Heterogeneity of Treatment Effect Analyses

There was no evidence of a treatment effect with fluvoxamine as compared with placebo for vaccine status, timing of symptom onset, severity of symptoms, age, sex, or calendar time. There was evidence for possible differential treatment effect on time to recovery for BMI (heterogeneity of treatment effect p=0.014), with a suggestion of increasing treatment benefit with increasing BMI (**eFigure 4**).

### Adverse Events

Among participants who reported taking study drug at least once, AEs were uncommon and similar in both groups (5.2% [32/615] with fluvoxamine vs. 6.2% [35/565] with placebo) (**eTable 2**).

## DISCUSSION

Among outpatients with mild to moderate COVID-19, treatment with fluvoxamine 50 mg twice daily for 10 days compared with placebo did not improve time to recovery in this large trial of 1288 participants. The lack of treatment effect was also seen for secondary outcomes including hospitalization, death, and acute care visits. Hospitalization and death were uncommon in our largely vaccinated study population. These findings do not support the use of fluvoxamine at this dose and duration in patients with mild to moderate COVID-19.

There are numerous conflicting trials for use of fluvoxamine, and some of the differences can be attributable to dosage.^7,8,10,15^ In comparison with the largest published trial to date, the TOGETHER trial, ACTIV-6 has some similarities and differences. The TOGETHER trial utilized a higher dose of fluvoxamine (100 mg twice daily) compared with 50 mg twice daily in ACTIV-6. The timing of the study was also different, as the TOGETHER trial reported randomization to fluvoxamine from January 20 to August 5, 2021, predating the arrival of the SARS-CoV-2 omicron variant and associated subvariants.^10^ By contrast, ACTIV-6 was conducted during the delta and omicron variant surges. Lastly, these trials enrolled different populations. The TOGETHER trial’s fluvoxamine participants were unvaccinated, high-risk, symptomatic Brazilian adults with a known risk factor for progression to severe COVID-19. ACTIV-6 enrolled adults in the United States regardless of COVID-19 risk factors or vaccination status. In fact, the majority (67%) of patients enrolled in ACTIV-6 reported at least 2 doses of a SARS-CoV-2 vaccine. Similar null results were observed in the 2022 COVID-Out trial, which also tested fluvoxamine 50 mg twice daily versus placebo in a majority vaccinated population with overweight or obesity.^15^

ACTIV-6 has several strengths. As a nationwide trial in the United States, ACTIV-6 is a generalizable trial for all adults aged ≥30 years old with COVID-19. This trial enrolled rapidly during the delta and omicron variant surges and included vaccinated patients, thus remaining a highly relevant population for the present times. ACTIV-6 also has limitations. Due to the broadly inclusive study population, few clinical events occurred, resulting in an inability to study the treatment effect on clinical outcomes such as hospitalization. Due to the remote nature of the trial, the median time from symptom onset to receipt of study drug was 5 days, which is at the upper limit of the recommend start of antiviral medicines (≤5 days). However, we did not observe any significant interaction with respect to the time from symptom onset to study drug receipt, consistent with other trials.^10,15^

The ACTIV-6 trial did not identify a clinically relevant treatment effect with fluvoxamine 50 mg twice daily for 10 days compared with placebo as outpatient COVID-19 treatment. We did not observe any faster time to clinical recovery in the population studied. Unlike prior randomized trials of fluvoxamine at 100 mg twice daily, we did not see any impact on prevention of clinical progression to severe COVID-19 with a lower dose of fluvoxamine.^8,10^

## Data Availability

Prior to deposition of the data in a public repository which will occur when the platform trial has concluded, persons may request the data by submitting a proposal. If the data can be used for the proposed purpose and it is consistent with the informed consent, then the data will be released under a Data Use Agreement.

## Acknowledgments

We thank Samuel Bozzette, MD, PhD, and Eugene Passamani, MD, both of NCATS, for their roles in the trial design and protocol development. We also thank the ACTIV-6 Data Monitoring Committee and Clinical Events Committee Members (listed below) for their contributions. Data Monitoring Committee: Clyde Yancy, MD, MSc, Northwestern University Feinberg School of Medicine; Adaora Adimora, MD, University of North Carolina, Chapel Hill; Susan Ellenberg, PhD, University of Pennsylvania; Kaleab Abebe, PhD, University of Pittsburgh; Arthur Kim, MD, Massachusetts General Hospital; John D. Lantos, MD, Children’s Mercy Hospital; Jennifer Silvey-Cason, Participant representative; Frank Rockhold, PhD, Duke Clinical Research Institute; Sean O’Brien, PhD, Duke Clinical Research Institute; Frank Harrell, PhD, Vanderbilt University Medical Center; Zhen Huang, MS, Duke Clinical Research Institute. Clinical Events Committee: Renato Lopes, MD, PhD, MHS, W. Schuyler Jones, MD, Antonio Gutierrez, MD, Robert Harrison, MD, David Kong, MD, Robert McGarrah, MD, Michelle Kelsey, MD, Konstantin Krychtiuk, MD, Vishal Rao, MD all of the Duke Clinical Research Institute, Duke University School of Medicine.

## Author Contributions

Drs Naggie and Hernandez had full access to all of the data in the study and take responsibility for the integrity of the data and the accuracy of the data analysis. Drs Lindsell and Stewart directly accessed and verified the underlying study data. All authors contributed to the drafting and review of the manuscript and agreed to submit for publication.

## Disclosures

**McCarthy:** Nothing to report.

**Naggie:** Reports grants from NIH, the sponsor for this study, during the conduct of the study; Institutional research grants from Gilead Sciences, AbbVie; Consulting fees from Pardes Biosciences; Scientific advisor/Stock options from Vir Biotechnology; Consulting with no financial payment from Silverback Therapeutics; DSMB fees from Personal Health Insights, Inc; Event adjudication committee fees from BMS/PRA outside the submitted work.

**Boulware:** Reports grants from NIH during the conduct of the study.

**Lindsell:** Reports institutional grants from NCATS during the conduct of the study; Institutional grants from NIH, CDC, and DoD; Contract with institution for research services from Endpoint Health, bioMerieux, Entegrion Inc, Abbvie, and Astra Zeneca outside the submitted work; Dr Lindsell has a patent for risk stratification in sepsis and septic shock issued to Cincinnati Children’s Hospital Medical Center.

**Stewart:** Reports grants from NIH NCATS during the conduct of the study; Grants from NIH outside the submitted work.

**Felker:** Reports institutional research grants from NIH during the conduct of the study and from Novartis outside the submitted work.

**Jayaweera:** Reports grants from NCATS PI-Ralph Sacco during the conduct of the study; Grants from Gilead, Pfizer, Janssen, and Viiv; Consulting fees from Theratechnologies outside the submitted work.

**Sulkowski:** Reports advisory board fees from AbbVie, Gilead, GSK, Atea, Antios, Precision Bio, Viiv, and Virion; Institutional grants from Janssen outside the submitted work.

**Gentile:** Reports personal fees from Duke University for protocol development and oversight during the conduct of the study; grants from NIH outside the submitted work

**Bramante:** Nothing to report.

**Singh:** Advisor to Gilead Sciences, Regeneron Pharmaceuticals, and Medscape/WebMD. **Dolor:** Reports institutional grant funding from NIA; honorarium from the following: PCORI as a member of the PCORI Clinical Trials Advisory Panel, Einstein-Montefiore as a member of their CTSA External Advisory Board, Veterans Administration as a consultant to the Women’s Health Practice-Based Research Network all outside the submitted work.

**Ruiz-Unger:** To be provided.

**Wilson:** Reports institutional research funding from the National Center for Advancing Translational Sciences (3U24TR001608) during the conduct of the study.

**DeLong:** Reports institutional research funding from the National Center for Advancing Translational Sciences (3U24TR001608) during the conduct of the study.

**Remaly:** Reports institutional research funding from the National Center for Advancing Translational Sciences (3U24TR001608) during the conduct of the study.

**Wilder:** Reports institutional research funding from the National Center for Advancing Translational Sciences (3U24TR001608) during the conduct of the study.

**Collins:** Reports grant funding from NHLBI and personal fees from Vir Biotechnology during the conduct of the study.

**Dunsmore:** Nothing to report.

**Adam:** Reports other from US Government Funding through OWS during the conduct of the study.

**Thicklin:** Nothing to report.

**Hanna:** Reports grants from US Biomedical Advanced Research & Development Authority contract to Tunnell Government Services for consulting services during the conduct of the study; Personal fees from Merck & Co. and AbPro outside the submitted work.

**Ginde:** Reports grants from NIH during the conduct of the study; Grants from NIH, CDC, DoD, AbbVie (investigator-initiated), and Faron Pharmaceuticals (investigator-initiated) outside the submitted work.

**Castro:** Reports institutional grant funding from NIH, ALA, PCORI, AstraZeneca, GSK, Novartis, Pulmatrix, Sanofi-Aventis, Shionogi; Speaker/Consultant fees from Grant Funding, Genentech, Teva, Sanofi-Aventis; Consultant fees from Merck, Novartis, Arrowhead, OM Pharma, Allakos; Speaker honorarium from Amgen, AstraZeneca, GSK, Regeneron; Royalties from Elsevier all outside the submitted work.

**McTigue:** Reports grants from NIH Research Subcontract to the University of Pittsburgh during the conduct of the study; Research contract to the University of Pittsburgh from Pfizer, and Janssen outside the submitted work.

**Shenkman:** Nothing to report.

**Hernandez:** Reports grants from American Regent, Amgen, Boehringer Ingelheim, Merck, Verily, Somologic, and Pfizer; Personal fees from AstraZeneca, Boston Scientific, Cytokinetics, Bristol Myers Squibb, and Merck outside the submitted work.

## Funding

ACTIV-6 is funded by the National Center for 10 Advancing Translational Sciences (NCATS) (3U24TR001608-06S1). Additional support for this study was provided by the Office of the Assistant Secretary for Preparedness and Response, Biomedical Advanced Research and Development Authority (contract No.75A50122C00037). The Vanderbilt University Medical Center CTSA Clinical and Translational Science Award from NCATS (UL1TR002243) supported the REDCap infrastructure. Additional support was provided by the Vanderbilt University Medical Center Recruitment Innovation Core (U24TR001579).

## Notes

### Competing Interest Statement

Competing interests for all authors are provided in the manuscript.

### Clinical Trial

NCT04885530

### Author Declarations

WCG IRB gave ethical approval for this work.

### Summary of Updates

The number of sites has been updated and the grant number has been updated.

