## Supplementary material for "Fluvoxamine for Outpatient Treatment of COVID-19: A Decentralized, Placebo-controlled, Randomized, Platform Clinical Trial": Online supplement

**Online-only Supplement**

### **ACTIV-6 Executive Committee**

Adrian F. Hernandez, Duke Clinical Research Institute (Clinical Coordinating Center PI)

Susanna Naggie, Duke Clinical Research Institute (Clinical Coordinating Center Co-PI)

G. Michael Felker, Duke Clinical Research Institute (Medical Monitor, blinded)

Sybil Wilson, Duke Clinical Research Institute

Allison DeLong, Duke Clinical Research Institute

April Remaly, Duke Clinical Research Institute

Rhonda Wilder, Duke Clinical Research Institute

Christopher J. Lindsell, Vanderbilt University Medical Center (Data Coordinating Center PI)

Thomas G. Stewart, Vanderbilt University Medical Center

Sean Collins, Vanderbilt University Medical Center

Sarah Dunsmore, National Center for Advancing Translational Sciences

Sam Bozzette, National Center for Advancing Translational Sciences

Gene Passamani, National Center for Advancing Translational Sciences

Stacey Adam, Foundation for the National Institutes of Health

David Boulware, University of Minnesota

Elizabeth Shenkman, University of Florida

Florence Thicklin, Stakeholder Advisory Committee

Matthew William McCarthy, Weill Cornell Medicine, Stakeholder Advisory Committee

George Hanna, Biomedical Advanced Research and Development Authority

### **ACTIV-6 Protocol Oversight Committee**

David Boulware, University of Minnesota, POC Co-Chair

Elizabeth Shenkman, University of Florida, POC Co-Chair

Adrian F. Hernandez, Duke Clinical Research Institute (Clinical Coordinating Center PI)

Susanna Naggie, Duke Clinical Research Institute (Clinical Coordinating Center co-PI)

G. Michael Felker, Duke Clinical Research Institute (Medical Monitor, blinded)

Sybil Wilson, Duke Clinical Research Institute

Allison DeLong, Duke Clinical Research Institute

April Remaly, Duke Clinical Research Institute

Rhonda Wilder, Duke Clinical Research Institute

Christopher J. Lindsell, Vanderbilt University Medical Center (Data Coordinating Center PI)

Thomas G. Stewart, Vanderbilt University Medical Center

Sean Collins, Vanderbilt University Medical Center

Sarah Dunsmore, National Center for Advancing Translational Sciences

Sam Bozzette, National Center for Advancing Translational Sciences

Gene Passamani, National Center for Advancing Translational Sciences

Stacey Adam, Foundation for the National Institutes of Health

Florence Thicklin, Stakeholder Advisory Committee

Matthew William McCarthy, Weill Cornell Medicine, Stakeholder Advisory Committee

George Hanna, Biomedical Advanced Research and Development Authority

Adit Ginde, University of Colorado Denver – Anschutz

Mario Castro, University of Kansas Medical Center

Dushyantha Jayaweera, University of Miami

Mark Sulkowski, John Hopkins University

Nina Gentile, Lewis Katz School of Medicine at Temple University

Kathleen McTigue, University of Pittsburgh Medical Center

Kim Marschhauser, PCORI

Julia Garcia-Diaz, Ochsner Health

### **ACTIV-6 Clinical Trial Team**

Adrian F. Hernandez, Duke Clinical Research Institute (Clinical Coordinating Center PI)

Susanna Naggie, Duke Clinical Research Institute (Clinical Coordinating Center Co-PI)

G. Michael Felker, Duke Clinical Research Institute (Medical Monitor, blinded)

Sybil Wilson, Duke Clinical Research Institute

Allison DeLong, Duke Clinical Research Institute

April Remaly, Duke Clinical Research Institute

Rhonda Wilder, Duke Clinical Research Institute

Christopher J. Lindsell, Vanderbilt University Medical Center (Data Coordinating Center PI)

Thomas G. Stewart, Vanderbilt University Medical Center

### **ACTIV-6 Independent Data Monitoring Committee**

Voting Members:

Clyde Yancy, Northwestern University Feinberg School of Medicine (Chair)

Adaora Adimora, University of North Carolina, Chapel Hill (Vice-chair)

Susan Ellenberg, University of Pennsylvania

Kaleab Abebe, University of Pittsburgh

Arthur Kim, Massachusetts General Hospital

John D. Lantos, Children’s Mercy Hospital

Jennifer Silvey-Cason, Participant representative

Statistical Data Analysis Center:

Frank Rockhold, Duke Clinical Research Institute (Lead faculty statistician)

Sean O’Brien, Duke Clinical Research Institute (Faculty statistician)

Frank Harrell, Vanderbilt University Medical Center (DCC-SDAC Faculty Liaison)

Zhen Huang, Duke Clinical Research Institute (Lead statistician)

### **ACTIV-6 Clinical Events Classification Committee**

Renato Lopes, (CEC PI)

Faculty Reviewers: W. Schuyler Jones, Antonio Gutierrez, Robert Harrison, David Kong, Robert McGarrah

Fellow Reviewers: Michelle Kelsey, Konstantin Krychtiuk, Vishal Rao

### **ACTIV-6 Data Coordinating Center**

David Aamodt, Vanderbilt University Medical Center

JaMario Ayers, Vanderbilt University Medical Center

Jess Collins, Vanderbilt University Medical Center

John Graves, Vanderbilt University Medical Center

James Grindstaff, Vanderbilt University Medical Center

Frank Harrell, Vanderbilt University Medical Center (DCC-SDAC Faculty Liaison)

Jessica Lai, Vanderbilt University Medical Center

Christopher J. Lindsell, Vanderbilt University Medical Center (Data Coordinating Center PI)

Itzel Lopez, Vanderbilt University Medical Center

Jessica Marlin, Vanderbilt University Medical Center

Alyssa Merkel, Vanderbilt University Medical Center

Sam Nwosu, Vanderbilt University Medical Center

Savannah Obregon, Vanderbilt University Medical Center

Dirk Orozco, Vanderbilt University Medical Center

Yoli Perez-Torres, Vanderbilt University Medical Center

Nelson Prato, Vanderbilt University Medical Center

Colleen Ratcliff, Vanderbilt University Medical Center

Max Rohde, Vanderbilt University Medical Center

Russell Rothman, Vanderbilt University Medical Center

Jana Shirey-Rice, Vanderbilt University Medical Center

Krista Vermillion, Vanderbilt University Medical Center

Thomas Stewart, Vanderbilt University Medical Center (Lead statistician)

Hsi-nien Tan, Vanderbilt University Medical Center

Seibert Tregoning, Vanderbilt University Medical Center

Meghan Vance, Vanderbilt University Medical Center

Amber Vongsamphanh, Vanderbilt University Medical Center

Maria Weir, Vanderbilt University Medical Center

Nicole Zaleski, Vanderbilt University Medical Center

### **ACTIV-6 Primary Site Investigators / Study Coordinators**

**A New Start II, LLC:** William (Kelly) Vincent / Raina Vincent. **Advanced Medical Care, Ltd:** Ray Bianchi / Jen Premas. **AMRON Vitality and Wellness Center, LLC:** Diana Cordero-Loperena / Evelyn Rivera. **Ananda Medical Clinic:** Madhu Gupta / Greg Karawan & Carey Ziomek. **Arena Medical Group:** Joseph Arena / Sonaly DeAlmeida. **Assuta Family Medical Group APMC:** Soroush Ramin / Jaya Nataraj. **Boston Medical Center:** Michael Paasche-Orlow / Lori Henault & Katie Waite. **Bucks County Clinical Research:** David Miller / Ginger Brounce. **Christ the King Health Care, P.C.:** Constance George-Adebayo / Adeolu Adebayo. **Clinical Trials Center of Middle Tennessee:** Alex Slandzicki / Jessica Wallan. **Comprehensive Pain Management and Endocrinology:** Claudia Vogel / Sebastian Munoz. **David Kavtaradze MD, Inc.:** David Kavtaradze / Cassandra Watson. **David Singleton MD, PA:** David Singleton / Maria Rivon & Amanda Sevier. **Del Pilar Medical and Urgent Care:** Arnold Del Pilar / Amber Spangler. **DHR Health Institute for Research:** Sohail Rao / Luis Cantu.

**Diabetes and Endocrinology Assoc. of Stark County:** Arvind Krishna / Kathy Evans, Tylene Falkner & Brandi Kerr. **Doctors Medical Group of Colorado Springs, P.C.:** Robert Spees / Mailyn Marta. **Duke University:** G. Michael Felker / Amanda Harrington. **Duke University Hospital:** Rowena Dolor / Madison Frazier, Lorraine Vergara & Jessica Wilson. **Elite Family Practice:** Valencia Burruss / Terri Hurst. **Emory University:** Igho Ofotokun / Laurel Bristow. **Essentia Health:** Rajesh Prabhu / Krystal Klicka & Amber Lightfeather. **Essential Medical Care, Inc.:** Vicki James / Marcella Rogers. **Express Family Clinic:** Pradeep Parihar / De'Ambra Torress. **Family Practice Doctors P.A.:** Chukwuemeka Oragwu / Ngozi Oguego. **First Care Medical Clinic:** Rajesh Pillai / Mustafa Juma. **Focus Clinical Research Solutions:** Ahab Gabriel / Emad Ghaly. **Franciscan Health Michigan City:** Dafer Al-Haddadin / Courtney Ramirez. **G&S Medical Associates, LLC:** Gammal Hassanien / Samah Ismail. **George Washington University Hospital:** Andrew Meltzer / Seamus Moran. **Geriatrics and Medical Associates:** Scott Brehaut / Angelina Roche. **GFC of Southeastern Michigan, PC:** Manisha Mehta / Nicole Koppinger. **Health Quality Primary Care:** Jose Baez / Ivone Pagan. **Highlands Medical Associates, P.A.:** Dallal Abdelsayed / Mina Aziz. **Hoag Memorial Hospital Presbyterian:** Philip Robinson / Julie Nguyen. **Hugo Medical Clinic:** Victoria Pardue / Llisa Hammons. **Innovation Clinical Trials Inc.:** Juan Ruiz-Unger / Susan Gonzalez & Lionel Reyes. **Jackson Memorial Hospital:** John Cienki / Gisselle Jimenez. **Jadestone Clinical Research, LLC:** Jonathan Cohen / Matthew Wong & Ying Yuan. **Jeremy W. Szeto, D.O., P.A.:** Jeremy Szeto. **Johns Hopkins Hospital:** Mark Sulkowski / Lauren Stelmash. **Lakeland Regional Medical Center:** Arch Amon / Daniel Haight. **Lamb Health, LLC:** Deryl Lamb / Amron Harper. **Lice Source Services Plantation:** Nancy Pyram-Bernard / Arlen Quintero. **Lupus Foundation of Gainesville:** Eftim Adhami. **Maria Medical Center, PLLC:** Josette Maria / Diksha Paudel & Oksana Raymond. **Medical Specialists of Knoxville:** Jeffrey Summers / Tammy Turner. **Medical University of South Carolina:** Leslie Lenert / Sam Gallegos & Elizabeth Ann Szwast. **Mediversity Healthcare:** Ahsan Abdulghani / Pravin Vasoya. **Miller Family Practice, LLC:** Conrad Miller / Hawa Wiley. **North Shore University Health System/Evanston Hospital:** Nirav Shah / Tovah Klein. **Ochsner Clinic Foundation:** Julie Castex / Phillip Feliciano. **Olivo Wellness Medical Center:** Jacqueline Olivo / Marian Ghaly, Zainub Javed & Alexandra Nawrocki. **Pine Ridge Family Medicine Inc.:** Anthony Vecchiarelli / Nikki Vigil. **Premier Health:** Vijaya Cherukuri / Erica Burden. **Rapha Family Wellness:** Dawn Linn / Laura Fisher. **Raritan Bay Primary Care & Cardiology Associates:** Vijay Patel / Praksha Patel & Yuti Patel.

**Romancare Health Services:** Leonard Ellison / Jeffrey Harrison. **Spinal Pain and Medical Rehab, PC:** Binod Shah / Sugata Shah. **Stanford University:** Upinder Singh / Julia Donahue & Yasmin Jazayeri. **Sunshine Walk In Clinic:** Anita Gupta / N Chandrasekar & Beth Moritz. **Tabitha B. Fortt, M.D., LLC:** Tabitha Fortt / Anisa Fortt. **Tallahassee Memorial Hospital:** Ingrid Jones-Ince / Alix McKee & Christy Schattinger. **Tampa General Hospital:** Jason Wilson / Brenda Farlow. **Temple University Hospital:** Nina Gentile / Lillian Finlaw. **Texas Health Physicians Group:** Randall Richwine & Tearani Williams / Penny Pazier & Lisa Carson. **Texas Tech University Health Sciences Center in El Paso:** Edward Michelson / Danielle Austin. **The Heart and Medical Center:** Sangeeta Khetpal / Tiffaney Cantrell, Drew Franklin & Karissa Marshall. **Trident Health Center:** Arvind Mahadevan / Madelyn Rosequist. **TriHealth, Inc:** Martin Gnoni / Crystal Daffner. **UF Health Precision Health Research:** Carla VandeWeerd / Mitchell Roberts. **University Diagnostics and Treatment Clinic:** Mark D'Andrea / Mina Aziz. **University Medical Center- New Orleans:** Stephen Lim / Wayne Swink. **University of Cincinnati:** Margaret Powers-Fletcher / Sylvere Mukunzi. **University of Florida Health:** Elizabeth Shenkman / Jamie Hensley & Brittney Manning. **University of Florida-JAX-ASCENT:** Carmen Isache / Jennifer Bowman, Angelique Callaghan-Brown & Taylor Scott. **University of Kansas – Wichita:** Tiffany Schwasinger-Schmidt / Ashlie Cornejo. **University of Miami:** Dushyantha Jayaweera / Maria Almanzar, Letty Ginsburg & Americo Hajaz. **University of Minnesota:** Carolyn Bramante. **University of Missouri – Columbia:** Matthew Robinson / Michelle Seithel. **University of Pittsburgh:** Akira Sekikawa / Emily Klawson. **University of Texas Health Science Center at Houston:** Luis Ostrosky / Virginia Umana. **University of Texas Health Science Center at San Antonio:** Thomas Patterson / Robin Tragus. **University of Virginia Health System:** Patrick Jackson / Caroline Hallowell & Heather Haughey. **Vaidya MD PLLC:** Bhavna Vaidya-Tank / Cameron Gould. **Vanderbilt University Medical Center:** Parul Goyal / Carly Gatewood. **Wake Forest University Health Sciences:** John Williamson / Hannah Seagle. **Weill Cornell Medical College:** Matthew McCarthy / Elizabeth Salsgiver. **Well Pharma Medical Research:** Eddie Armas / Jhonsai Cheng & Priscilla Huerta.

### **Supplemental Methods**

**Participant Monitoring**

The daily and follow-up assessments were monitored, and sites were actively notified of events requiring review, including serious adverse events. In addition, participants were invited during these assessments to request contact from the study team or to report any unusual circumstances that might be relevant. Failure to complete daily assessments was also a trigger for review of a possible serious adverse event. A missed assessment on the day after receiving the first dose of study medication (day 2) or any day of missed assessments up to day 14 prompted an investigator notification to contact the participant. All participants were instructed to self-report concerns either via an online event reporting system, by calling the site, or by calling a 24-hour hotline.

Hospitalizations, a record of seeking other healthcare, or serious adverse events were extracted by site personnel from the participant’s medical record. Medical occurrences occurring before the receipt of study drug/placebo but after obtaining informed consent were not considered an adverse event.

**Independent Data Monitoring Committee Oversight**

Interim analyses were planned at intervals of approximately 300 participants contributing to a study drug group, with an anticipated maximum of 1200 participants. There was also the potential to extend accrual for a study drug if there was potential to demonstrate benefit for hospitalization/death. Due to extremely rapid enrollment related to the omicron variant surge, the first planned interim analysis was not conducted. The independent data monitoring committee reviewed interim data when approximately 900 participants were enrolled only, resulting in a planned primary analysis highly conservative of type 1 error. To provide additional context, the primary analysis was additionally performed with a non-informative prior and without a prior.

**Handling of Missing Data**

In both the primary and secondary endpoint analyses, missing data among covariates was addressed with conditional mean imputation because the amount of missing covariate data was small. Approximately 7–8% of participants did not report activity level for the COVID clinical progression score endpoint at each time point, but the participants were known to be alive and at home. The missing activity level was a type of interval censored outcome, as the participants were known to be either a 1 or 2 on the scale. The ordinal regression models were fit accounting for the interval censoring. The proportional hazards assumption of the primary endpoint was evaluated by generating visual diagnostics such as the log-log plot and plots of time-dependent regression coefficients for each predictor in the model, a diagnostic which indicates deviations from proportionality if the time-dependent coefficients are not constant in time.

**Heterogeneity of Treatment Effect Analysis**

For each characteristic, a proportional hazards regression model was constructed using the same covariates as the primary endpoint model plus additional interaction terms between treatment assignment and the characteristic of interest. To allow the possibility of non-linear trends along continuous characteristics, like age or calendar time, continuous covariates were included in the model as restricted cubic splines. The hazard ratios and 95% confidence intervals were calculated from asymptotic, model-based estimates at specific values. The continuous variables were not discretized into bins (or groups).

### **COVID-19 Ordinal Outcome Scale**

The COVID-19 outcomes for this trial are based on the World Health Organization’s Ordinal Scale for Clinical Improvement and will be collected via the online system and from the medical record. The following outcomes will be assessed as part of the COVID Clinical Progression Scale:

0. No clinical or virological evidence of infection

1. No limitation of activities

2. Limitation of activities

3. Hospitalized, no oxygen therapy

4. Hospitalized, on oxygen by mask or nasal prongs

5. Hospitalized, on non-invasive ventilation or high-flow oxygen

6. Hospitalized, on intubation and mechanical ventilation

7. Hospitalized, on ventilation + additional organ support – pressors, RRT, ECMO

8. Death

### **eTable 1. Baseline symptom prevalence and severity**

| **Variable** | **Fluvoxamine** | **Placebo** | **Overall** |
| --- | --- | --- | --- |
|  | **(n=674)** | **(n=614)** | **(n=1288)** |
| Symptom burden on study day 1, No./total (%) |  |  |  |
| None | 36/622 (5.79) | 37/569 (6.50) | 73/1191 (6.13) |
| Mild | 396/622 (63.67) | 353/569 (62.04) | 749/1191 (62.89) |
| Moderate | 176/622 (28.30) | 166/569 (29.17) | 342/1191 (28.72) |
| Severe | 14/622 (2.25) | 13/569 (2.28) | 27/1191 (2.27) |
| Symptoms on study day 1 |  |  |  |
| Fatigue, No./total (%) |  |  |  |
| None | 60/586 (10.24) | 43/533 (8.07) | 103/1119 (9.20) |
| Mild | 308/586 (52.56) | 290/533 (54.41) | 598/1119 (53.44) |
| Moderate | 197/586 (33.62) | 172/533 (32.27) | 369/1119 (32.98) |
| Severe | 21/586 (3.58) | 28/533 (5.25) | 49/1119 (4.38) |
| Dyspnea, No./total (%) |  |  |  |
| None | 313/586 (53.41) | 296/533 (55.53) | 609/1119 (54.42) |
| Mild | 212/586 (36.18) | 191/533 (35.83) | 403/1119 (36.01) |
| Moderate | 55/586 (9.39) | 43/533 (8.07) | 98/1119 (8.76) |
| Severe | 6/586 (1.02) | 3/533 (0.56) | 9/1119 (0.80) |
| Fever, No./total (%) |  |  |  |
| None | 446/586 (76.11) | 427/532 (80.26) | 873/1118 (78.09) |
| Mild | 108/586 (18.43) | 88/532 (16.54) | 196/1118 (17.53) |
| Moderate | 27/586 (4.61) | 15/532 (2.82) | 42/1118 (3.76) |
| Severe | 5/586 (0.85) | 2/532 (0.38) | 7/1118 (0.63) |
| Cough, No./total (%) |  |  |  |
| None | 88/586 (15.02) | 72/532 (13.53) | 160/1118 (14.31) |
| Mild | 325/586 (55.46) | 294/532 (55.26) | 619/1118 (55.37) |
| Moderate | 145/586 (24.74) | 146/532 (27.44) | 291/1118 (26.03) |
| Severe | 28/586 (4.78) | 20/532 (3.76) | 48/1118 (4.29) |
| Nausea, No./total (%) |  |  |  |
| None | 449/586 (76.62) | 415/532 (78.01) | 864/1118 (77.28) |
| Mild | 106/586 (18.09) | 87/532 (16.35) | 193/1118 (17.26) |
| Moderate | 28/586 (4.78) | 25/532 (4.70) | 53/1118 (4.74) |
| Severe | 3/586 (0.51) | 5/532 (0.94) | 8/1118 (0.72) |
| Vomiting, No./total (%) |  |  |  |
| None | 556/586 (94.88) | 509/532 (95.68) | 1065/1118 (95.26) |
| Mild | 20/586 (3.41) | 17/532 (3.20) | 37/1118 (3.31) |
| Moderate | 9/586 (1.54) | 6/532 (1.13) | 15/1118 (1.34) |
| Severe | 1/586 (0.17) | 0/532 (0.00) | 1/1118 (0.09) |
| Diarrhea, No./total (%) |  |  |  |
| None | 445/586 (75.94) | 402/532 (75.56) | 847/1118 (75.76) |
| Mild | 109/586 (18.60) | 109/532 (20.49) | 218/1118 (19.50) |
| Moderate | 28/586 (4.78) | 19/532 (3.57) | 47/1118 (4.20) |
| Severe | 4/586 (0.68) | 2/532 (0.38) | 6/1118 (0.54) |
| Body aches, No./total (%) |  |  |  |
| None | 189/586 (32.25) | 188/532 (35.34) | 377/1118 (33.72) |
| Mild | 285/586 (48.63) | 222/532 (41.73) | 507/1118 (45.35) |
| Moderate | 92/586 (15.70) | 101/532 (18.98) | 193/1118 (17.26) |
| Severe | 20/586 (3.41) | 21/532 (3.95) | 41/1118 (3.67) |
| Sore throat, No./total (%) |  |  |  |
| None | 255/586 (43.52) | 254/532 (47.74) | 509/1118 (45.53) |
| Mild | 238/586 (40.61) | 196/532 (36.84) | 434/1118 (38.82) |
| Moderate | 75/586 (12.80) | 66/532 (12.41) | 141/1118 (12.61) |
| Severe | 18/586 (3.07) | 16/532 (3.01) | 34/1118 (3.04) |
| Headache, No./total (%) |  |  |  |
| None | 224/586 (38.23) | 226/532 (42.48) | 450/1118 (40.25) |
| Mild | 236/586 (40.27) | 214/532 (40.23) | 450/1118 (40.25) |
| Moderate | 111/586 (18.94) | 73/532 (13.72) | 184/1118 (16.46) |
| Severe | 15/586 (2.56) | 19/532 (3.57) | 34/1118 (3.04) |
| Chills, No./total (%) |  |  |  |
| None | 420/586 (71.67) | 393/532 (73.87) | 813/1118 (72.72) |
| Mild | 125/586 (21.33) | 105/532 (19.74) | 230/1118 (20.57) |
| Moderate | 32/586 (5.46) | 30/532 (5.64) | 62/1118 (5.55) |
| Severe | 9/586 (1.54) | 4/532 (0.75) | 13/1118 (1.16) |
| Nasal symptoms, No./total (%) |  |  |  |
| None | 163/586 (27.82) | 137/532 (25.75) | 300/1118 (26.83) |
| Mild | 287/586 (48.98) | 277/532 (52.07) | 564/1118 (50.45) |
| Moderate | 115/586 (19.62) | 102/532 (19.17) | 217/1118 (19.41) |
| Severe | 21/586 (3.58) | 16/532 (3.01) | 37/1118 (3.31) |
| New loss of sense of taste or smell, No./total (%) |  |  |  |
| None | 354/586 (60.41) | 339/532 (63.72) | 693/1118 (61.99) |
| Mild | 116/586 (19.80) | 86/532 (16.17) | 202/1118 (18.07) |
| Moderate | 61/586 (10.41) | 62/532 (11.65) | 123/1118 (11.00) |
| Severe | 55/586 (9.39) | 45/532 (8.46) | 100/1118 (8.94) |

### **eTable 2. Adverse events**

| **Variable** | **Fluvoxamine, not taken** | **Fluvoxamine, taken** | **Placebo, not taken** | **Placebo, taken** | **Overall** |
| --- | --- | --- | --- | --- | --- |
|  | 59 | 615 | 49 | 565 | 1288 |
| Experienced an adverse events, No./total (%) | 0/55 (0.00) | 29 (4.72) | 1/42 (2.38) | 30 (5.31) | 60/1277 (4.70) |
| Experienced a serious adverse events, No./total (%) | 0/55 (0.00) | 3 (0.49) | 0/42 (0.00) | 5 (0.88) | 8/1277 (0.63) |
| Serious adverse events |  |  |  |  |  |
| COPD exacerbation |  | 2 |  | 0 | 2 |
| Broken ankle |  | 0 |  | 1 | 1 |
| Chest pain |  | 0 |  | 1 | 1 |
| Coronary vasospasm |  | 0 |  | 1 | 1 |
| Infected finger |  | 1 |  | 0 | 1 |
| Ophthalmic migraine |  | 0 |  | 1 | 1 |
| COVID-19 pneumonia |  | 0 |  | 1 | 1 |
| Note: “Taken” refers to the participants who reported taking (or planning to take) the study drug at least once. “Not taken” refers to the participants (if any) who did not report taking the study drug. | | | | | |

### **eFigure 1. Time from symptom onset to receipt of drug**

| 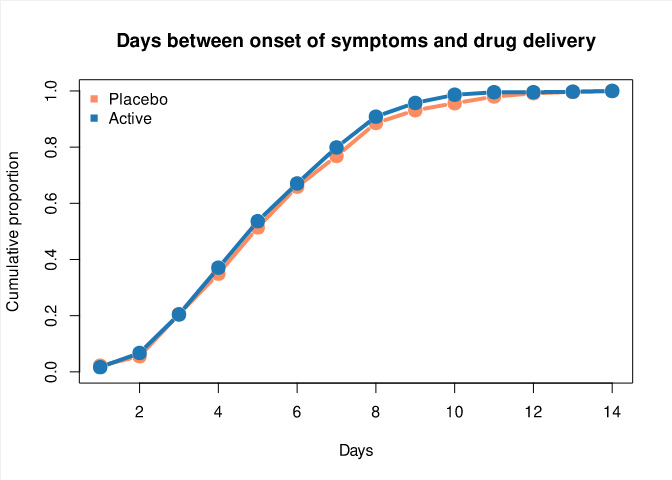 | | | | | | | | | | | | | | |
| --- | --- | --- | --- | --- | --- | --- | --- | --- | --- | --- | --- | --- | --- | --- |
| **Cumulative proportion by day** | | | | | | | | | | | | | | |
|  | **1** | **2** | **3** | **4** | **5** | **6** | **7** | **8** | **9** | **10** | **11** | **12** | **13** | **14** |
| Placebo | 0.02 | 0.06 | 0.21 | 0.35 | 0.51 | 0.66 | 0.77 | 0.89 | 0.93 | 0.96 | 0.98 | 0.99 | 1 | 1 |
| Active | 0.02 | 0.07 | 0.20 | 0.37 | 0.54 | 0.67 | 0.80 | 0.91 | 0.96 | 0.99 | 1.00 | 1.00 | 1 | 1 |

### **eFigure 2A. All-cause hospitalization or death for fluvoxamine versus placebo**

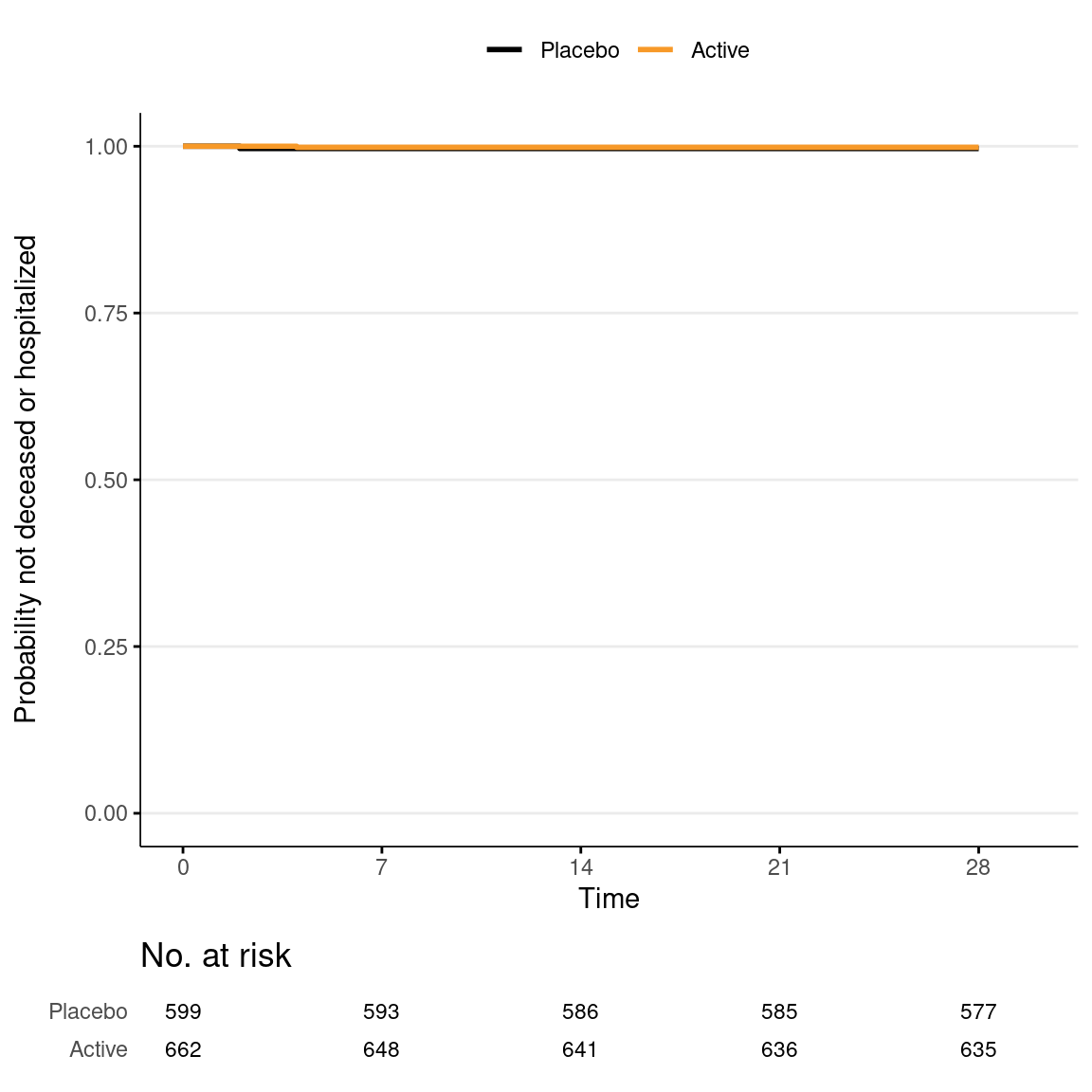

### **eFigure 2B. All-cause hospitalization, urgent care, emergency room visit, or death for fluvoxamine versus placebo**

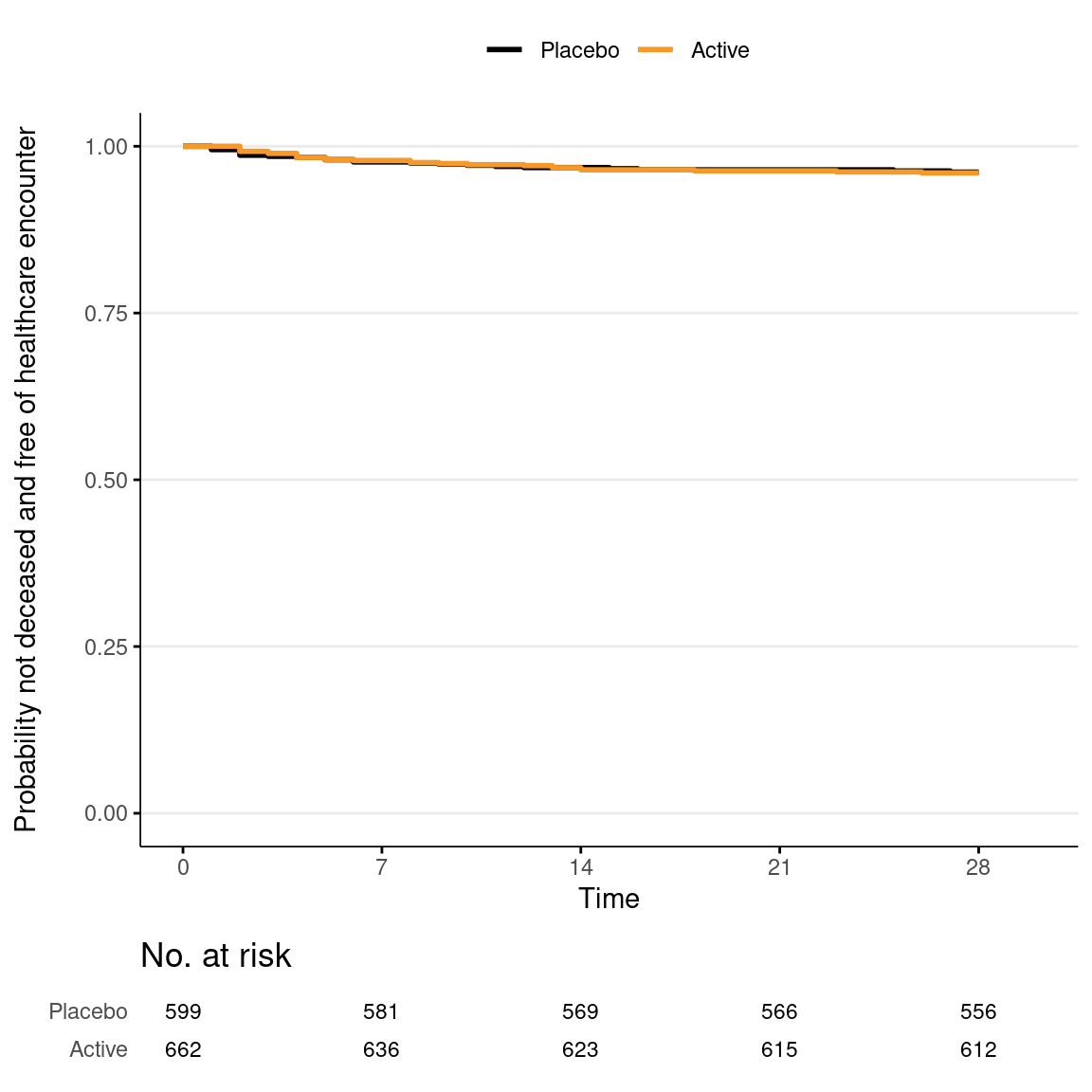

### **eFigure 3. Participants’ clinical status at day 7, 14, and 28**

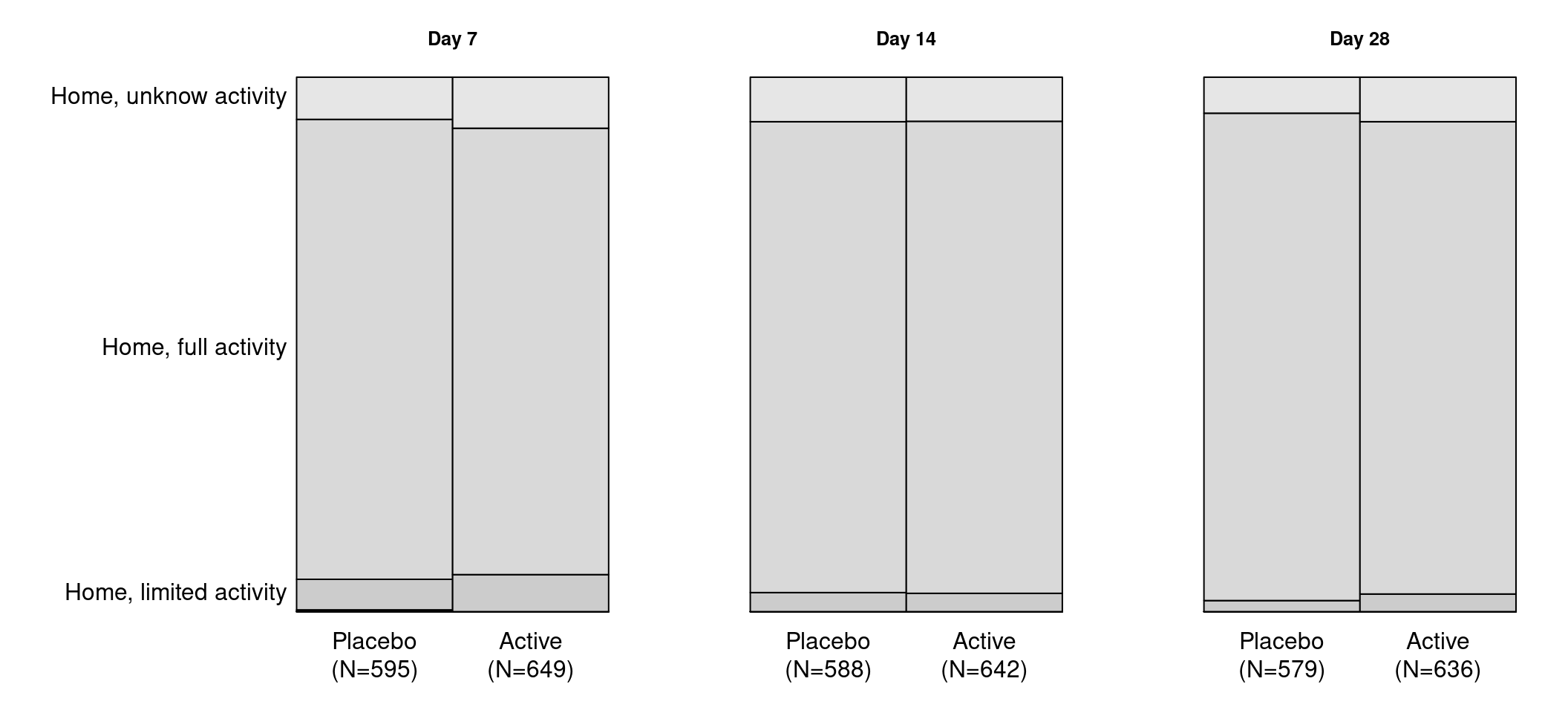

### **eFigure 4. Heterogeneity of treatment effect between fluvoxamine and placebo for time to recovery**

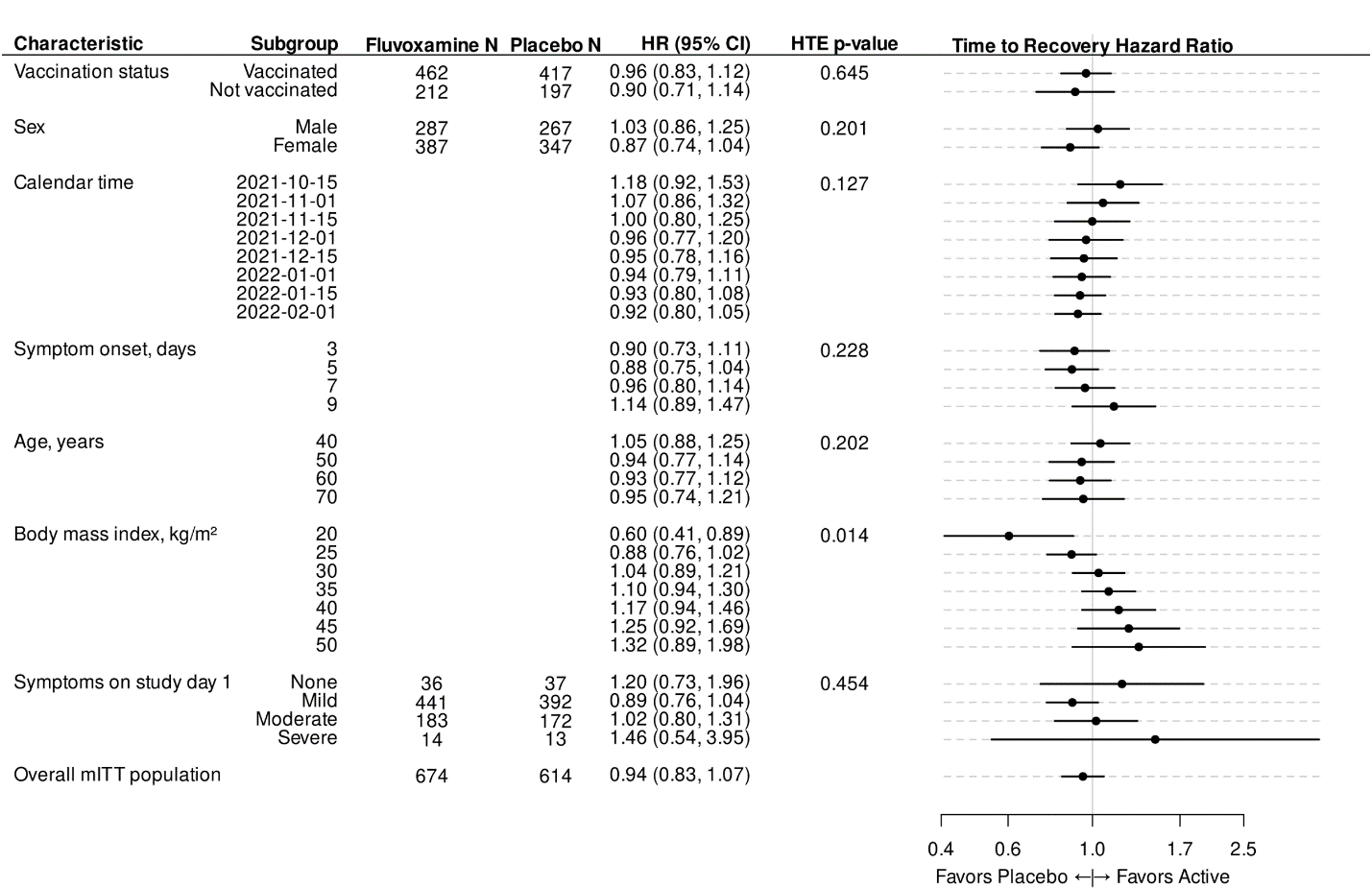
